# A questionnaire for a conceptual framework and interdisciplinary public health research using the Delphi technique – development and validation

**DOI:** 10.1101/2024.12.13.24318025

**Authors:** Anita Alaze, Emily Finne, Stephanie Batram-Zaantvoort, Oliver Razum, Céline Miani

**Affiliations:** Department of Epidemiology and International Public Health, School of Public Health, Bielefeld University, Universitätsstr. 25, 33615 Bielefeld, Germany

**Keywords:** questionnaire development, Delphi survey, interdisciplinarity, public health, conceptual framework

## Abstract

**Background:** The Delphi technique has become established in public health research, yet there is a lack of methodological standards in questionnaire development. We here demonstrate how the Delphi technique can be used in an interdisciplinary public health topic for framework development, and we highlight methodological challenges and possible solutions.

**Methods:** We developed the questionnaire through a comprehensive literature review and the generation of an item pool based on the rules of item construction. We used cognitive interviews, Delphi experts’ assessment and group discussions to refine the questionnaire and to ensure content validity. Finally, we pre-tested the online questionnaire on LimeSurvey.

**Results:** The questionnaire consists of three main sections, namely gender (norms), the social environment and the mental health of adolescents, and another section on the expert’s characteristics. The questionnaire comprises a total of 32 questions and includes rating and ranking questions, content-related and comment questions, open and closed questions as well as questions on personal characteristics and evaluation questions.

**Conclusion:** Interdisciplinary researchers need to be involved in the development process of the questionnaire to address challenges related to interdisciplinarity. Researchers using the Delphi technique should consider certain aspects of the development process, such as comprehensively preprocessing the content, disentangling the theoretical concepts, reducing the complexity, providing more structure and clarity, and ensuring that the length of the questionnaire is manageable. Future research should focus on developing methodological guidelines and testing their applicability for different objectives of the Delphi technique (e.g. framework development).

## Background

### Delphi technique

The Delphi technique, an anonymous expert-based assessment process carried out in a group communication process, is used by researchers if the available knowledge on a topic is uncertain or incomplete. The procedure is as follows. The topic under investigation is evaluated by experts in an iterative process (1). A Delphi study consists of several (2-4) rounds until a consensus is reached, ideas are aggregated, future predictions are made or experts’ opinions are determined (2). After each round, the answers are summarized and attached to each new questionnaire so that the experts surveyed can reconsider their opinions and revise them if necessary (2). The Delphi survey, compared to traditional group discussion, has numerous advantages, such as flexibility, limiting dominance of certain individuals, limiting moderator biases, enabling collection of opinions from participants located in various geographic regions and at the same time ensuring anonymity during the survey (3).

The objective for using the Delphi technique differs between disciplines. In public health, it is commonly used to find expert consensus. While the Delphi technique has gained acceptance in diverse fields of medicine and public health, little methodological guidance and quality standards are provided in the context of theory development (4). Thus, Delphi can be considered a technique rather than a fixed method. This leads to a lack of clarity on best practice in the application of the Delphi technique for framework development and even more so with an interdisciplinary and international expert panel. A systematic review revealed that only one single out of 80 included studies provided the Delphi questionnaires in the appendix (5). It also shows that many essential elements of the Delphi technique are not reported in the studies (5), which indicates the need for improving the use and the reporting of the Delphi technique.

### Gender norms, social environment, and mental health of adolescents

Adolescents experience sex differences in morbidity and mortality. Key determinants to explain these differences are socially constructed gender norms. While gender inequalities affect the lives of both boys and girls, they tend to disadvantage girls in several ways, for instance, unequal access to resources, power, education and discriminatory socio-cultural practices (6). These gender norms shape the way adolescents interact, form relationships, and engage in sexual and reproductive practices as well as most social behaviours (6). Gender norms are thus important to investigate, as they affect everyone, influence their health-related behaviours very early in life (around the age of two (8)) and can shape the whole life course. However, little is known about the factors that influence young adolescents’ gender norms and behaviours in relation to (mental) health (6).

Gender theory highlights the importance of a multidimensional *social environment* shaping the various gender norms and attitudes of individuals. Structural dimensions, such as institutional laws and policies as well as social structure and individual aspects are often mentioned as influential factors in the macro, meso- and micro-environment associated with health (9–12). Adolescents’ social environment determines how adolescents learn about and construct gendered attitudes and norms which in turn affect health (13). This is why shaping these norms is particularly important.

Differences in *mental health* status can be observed very early in life, and childhood and adolescence are critical periods of health promotion, as more than half of mental health disorders already start at these stages, many of them affecting adulthood (14). Rates of common mental health problems, such as depression or anxiety rise sharply in adolescence and peak in young adulthood (15). Depression, anxiety and behavioural disorders are among the leading causes of illness and disability, and suicide is the fourth leading cause of death among adolescents (16). Despite the multidimensional nature of this global challenge, many interventions persist in seeking solutions that only tackle the individual level (14).

### This study

This study aims at demonstrating how the Delphi technique can be used in an interdisciplinary public health topic for framework development which can be used in quantitative research, contributing to providing a base for more theory-driven research. Moreover, this article highlights methodological challenges and what has proven successful in framework development with a Delphi survey with international experts and derives practical recommendations for other studies.

We will illustrate this using the example of the development of a questionnaire for a Delphi study. The objective of the questionnaire is to analyse how a) gender norms, b) the social environment and c) the mental health of adolescents are theoretically connected and which are the most important dimensions or influencing factors for investigating those connections. There is clearly a need for a conceptual framework to map the interplay of the three topics under investigation. However, so far there is no such theoretical approach and also none that applies it to the age group of adolescents.

## Method

### Questionnaire development

Decisions for the whole Delphi study, which includes three Delphi rounds, methodologically influenced the questionnaire development for the first Delphi round. We decided to include experts from the Global North and South due to the global character of the research topic. However, this means that the questionnaire must anticipate and address a variety of language, cultural and knowledge differences. Further, we chose to include researchers as well as practitioners at the policy and implementation level (e.g. NGO members) in the Delphi survey to gain analytical perspectives. Another essential element is the inclusion of experts from different disciplines (e.g. gender experts, gender socialisation experts, mental health experts, experts in adolescence and multiple combinations of those) to address the intersections of the questionnaire topics. However, not many experts have specialised knowledge in multiple scientific fields.

The questionnaire development process (see Fig. 1) comprised 1) a comprehensive literature review to identify the content domain and theories, models or conceptual/theoretical frameworks within the research topics gender norms, social environment, and the mental health of adolescents, 2) the generation of a pool of instrument items based on the literature review and on the first author’s expertise and 3) the first set-up of the questionnaire. We generated all items according to the rules of item construction (comprehensive, positive, short, easy to understand, uniqueness, no universal expressions, no suggestive questions, no redundancy, closed questions to increase economy and objectivity of analysis) (17) and for the use in quantitative research. In addition, we adapted the items to typical response scales of Delphi surveys, such as the rating and ranking scales. After the first set-up of the questionnaire, we held 4) several cognitive interviews with one content expert per topic to refine the questionnaire and to ensure content validity (18). 5) Two Delphi experts then assessed the questionnaire for its appropriateness as a Delphi survey based on their experience with other Delphi studies and essential elements of the Delphi technique (e.g. length of questionnaire, type and formulation of questions and answer options, fit of study objective with selection criteria (validity, feasibility, importance, agreement or reliability), overall outlay of the Delphi study, definition of consensus). We implemented the resulting suggestions for improvement, item prioritisation and reduction through group discussions with content experts. We conducted 6) another cognitive interview with one psychology practitioner to assess the feasibility of the questionnaire for practitioners. 7) We implemented all findings in the questionnaire and 8) set them up on LimeSurvey. 9) Ultimately, we pre-tested the online questionnaire on LimeSurvey.

**Figure 1:**
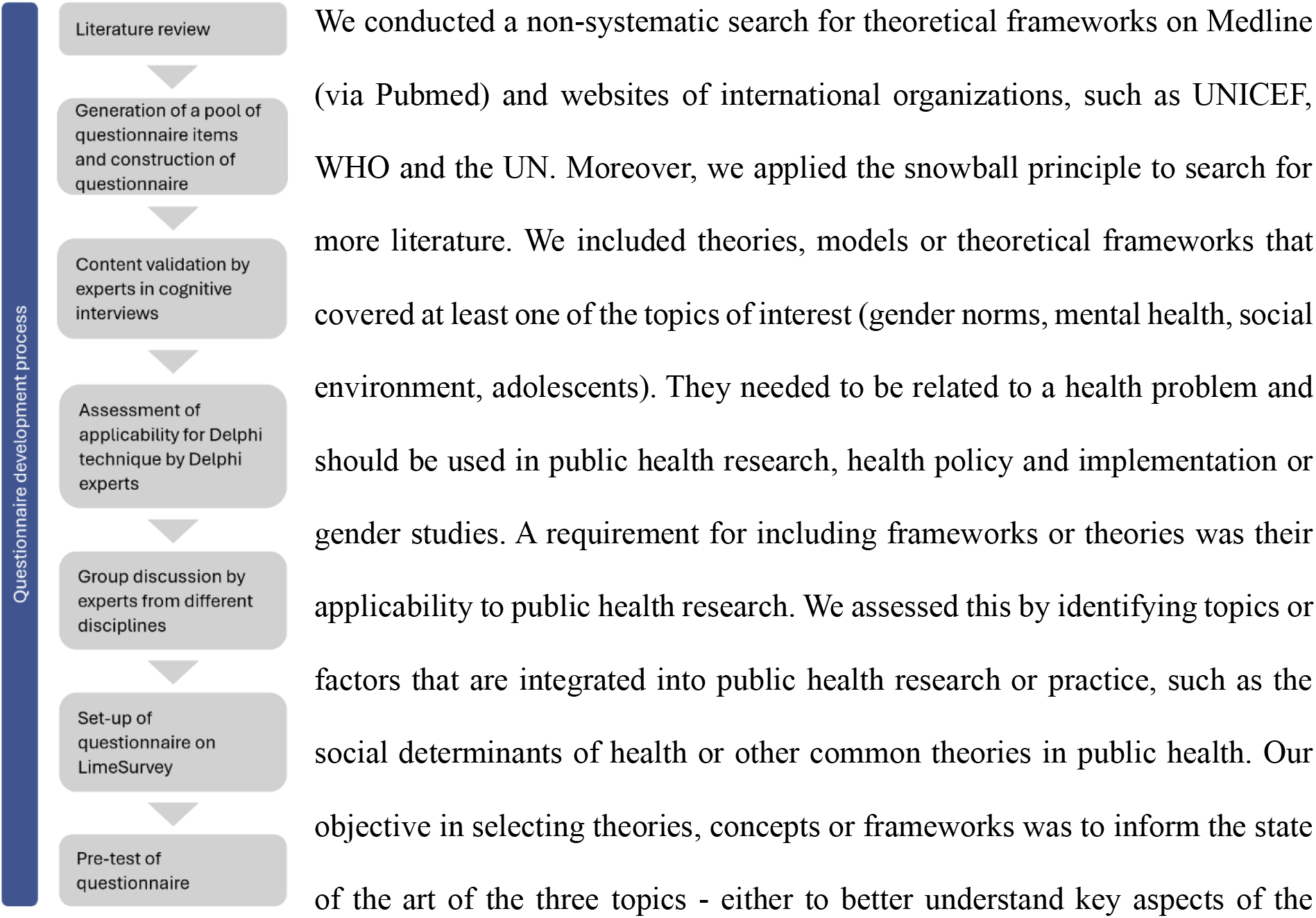
Flowchart of the development process of the Delphi questionnaire

### Literature review

We conducted a non-systematic search for theoretical frameworks on Medline (via Pubmed) and websites of international organizations, such as UNICEF, WHO and the UN. Moreover, we applied the snowball principle to search for more literature. We included theories, models or theoretical frameworks that covered at least one of the topics of interest (gender norms, mental health, social environment, adolescents). They needed to be related to a health problem and should be used in public health research, health policy and implementation or gender studies. A requirement for including frameworks or theories was their applicability to public health research. We assessed this by identifying topics or factors that are integrated into public health research or practice, such as the social determinants of health or other common theories in public health. Our objective in selecting theories, concepts or frameworks was to inform the state of the art of the three topics - either to better understand key aspects of the specific topic or to find connections between them. This resulted in the generation of an item pool and the first draft of the questionnaire.

### Expert interviews

#### Cognitive interviews

We carried out the cognitive interviews to ensure that each topic in the questionnaire covers the relevant content and to assess the understandability of the questions for experts, also in the areas where they do not have expertise. This is an important step to address the interdisciplinarity of the questionnaire during the development phase.

We held the first round of cognitive interviews after the first set-up of the questionnaire items with three scientists with expertise in at least one of the three topics. Then, we conducted the second round of cognitive interviews with a child and youth psychologist after the group discussion (step 6) to assess the understandability of the questionnaire by practitioners. The procedure was as follows: The experts received the questionnaire by email to think about the questionnaire in advance. During the interview (mostly via Zoom), we reviewed the questionnaire from top to bottom and asked the experts to comment on the understandability, completeness and relevance of the selected questionnaire items and accompanying response formats and to indicate missing or redundant items (content validity). We directed emphasis on the part of the questionnaire where the respondent has their expertise.

#### Delphi experts

We contacted two Delphi experts via email to assess if the questionnaire is applicable to and appropriate for the Delphi technique and if the research objective (to develop a conceptual framework with a Delphi study in an interdisciplinary research field) can be reached. Ultimately, we adapted the questionnaire based on the expert’s comments, feedback and suggestions to fit the Delphi technique, which we achieved through a group discussion. Notwithstanding, there is no set standard for the Delphi technique, making it difficult to know what exactly is required for a questionnaire to be suitable for the Delphi method.

#### Group discussion

We held group discussions to reduce the number of questionnaire items and answer options, to increase the clarity, to reduce the cognitive load and to adapt the layout and design to make it more appealing. We chose two scientists with a background in sociology and psychology who are currently working as public health researchers. This allowed us to address the interdisciplinarity of and the intersections between the research topics. We sent the questionnaire to the two experts before the group discussions. Ultimately, the group discussions included two meetings which we held in person. A challenge during the discussions was to decide which aspects should be predetermined (e.g. the most essential dimensions or definitions) to shorten the length of the questionnaire and which should be left as response options.

#### Pretest of the questionnaire

We pretested the questionnaire with researchers from the School of Public Health at Bielefeld University to check for spelling mistakes, layout, practicability, introductory text, ambiguities in questionnaire items, and set-up in LimeSurvey, an online survey program hosted on a server at Bielefeld University. Since it is challenging to convince busy experts to participate in a Delphi study with several rounds, we decided to carry out the pre-test within the faculty to maintain the chance of a higher participation rate in the actual survey.

## Results

### Literature review

We retrieved four relevant frameworks for the sex/gender concept and seven frameworks for gender and adolescence (see Tab. 1).

**Table 1:** Included frameworks in the literature review, grouped into frameworks addressing sex/gender concepts and the topic of gender and adolescence.

| Sex/gender concept | Gender and adolescence |
| --- | --- |
| INGER (Integrating Gender into Environmental health Research) multidimensional sex/gender concept with an intersectional perspective (12) | Ecological model for an enabling environment for shaping adolescent sexual and reproductive health (19) |
| The gender analysis framework (10) | GAGE's conceptual Framework (20) |
| Dynamic framework for social change (11) | Multi-level framework of influence impacting gender socialisation processes during adolescence (21) |
| Conceptual framework for structural elements of gender power relations that drive gender inequality (22) | Conceptual framework of social and gender norms and power for ASRH (Adolescent Sexual and Reproductive Health) (23) |
|  | Integrated model of menstrual experience (24) |
|  | Conceptual measurement framework for impacts of gender inequality on the well-being of children and adolescents (25) |
|  | Conceptual framework for healthy adolescent sexuality development and its potential link with sexual well-being (9) |

No framework addressed specifically gender and mental health. The general objectives of the frameworks were to inform research, develop interventions for practice or theoretically frame a research project concerning gender norms and adolescence. The literature review revealed that the interplay of these three topics is not or only partly covered in the existing literature. No framework, concept or model addressed the specific conceptualization of gender norms and even less so for adolescents. The social environment leading to the integration of the multi-level approach plays an important role in the frameworks, however, the specific inter-relational factors between the environmental levels and gender norms and the adolescent’s internalization process were not illustrated. In addition, mental health topics were rarely mentioned in conjunction with the other two topics, and when they were, it was only broadly acknowledged. Consequently, there is still a need to justify and clarify the use of these theoretical groundings to advance gender theoretical development (26). Nonetheless, those parts of the existing theories/concepts/frameworks that address one or more of the three constructs (gender (norms), the social environment or the mental health of adolescents) more specifically were integrated as questionnaire items or answer options into the conceptual framework for this research project.

We integrated diverse lenses or perspectives of the frameworks, such as intersectionality, embodiment, or power relations lens. These lenses derive from the state of the art and ‘gold standards’ in gender research. For instance, Hammarström and Hensing (2018) identified the six gender theoretical concepts of sex, gender, intersectionality, embodiment, gender equity and gender equality as central to health science (27). We included those concepts in the questionnaire to adhere to this standard. The frameworks served as examples as to how the concepts can be reflected in the framework to be developed.

#### Cognitive interviews, Delphi experts and group discussions

The cognitive interviews revealed which items of the questionnaire were incomprehensible. In addition, they indicated the necessity to include additional answer options. We restructured some sections to gain more clarity.

The evaluation of the questionnaire by the Delphi experts about its applicability to the Delphi technique revealed that the questionnaire should be further shortened to approximately 8 pages, the cognitive demand of the questions should be further reduced, just like the number of possible answers and open questions (e.g. one comment question per question and not per answer option). Moreover, experts advised to reduce the amount of explanation texts since they might not be read. Thus, the questions must contain all relevant information to answer the questions.

The qualitative group discussions further developed the questionnaire items towards more clarity, to reduce the cognitive load, and to adapt the design/layout aspects to make it more appealing to ultimately improve its applicability for the Delphi study. An obstacle towards greater clarity and a lower cognitive load was the many different theories from different disciplines which contain very similar concepts with different names and the question of how they can be combined into meaningful and distinguishable concepts. Therefore, we disentangled different gender/gender norms terms, further reduced the number of answer options as well as the comment sections (per section instead of per question) and implemented text boxes for definitions of terms. Where necessary, we have also adapted definitions to reflect the disentangled concepts and revised them to the most up-to-date understanding. Moreover, we introduced rating and/or hierarchical questions instead of asking to select the 3 most relevant aspects as we had done before.

#### Pretest of the questionnaire

The pretest revealed ambiguities in the questionnaire items, and the need for a better structured introductory text so that necessary information about the survey can be seen without reading the entire text, as well as technical aspects of the online survey.

**Table 2:** Constructs, indicators and indices of the core components of the Delphi questionnaire.

| Construct | Indicator | Indices |
| --- | --- | --- |
| Gender | Gender concepts | Current sex/gender identity<br>Sex/gender roles<br>Sex/gender expressions<br>Sex/gender relations<br>Sexuality (Sexual orientation) |
|  | Gender approaches | Gender continuum<br>Multidimensionality approach<br>Multi-level approach<br>Intersectionality approach<br>Gender power relations<br>Embodiment approach<br>Decolonial lens |
|  | Gender norms | General gender norms<br>Gender norms linked to mental health |
| Mental health | Outcomes for adolescent mental health potentially affected by gender norms | Mental, social and physical well-being<br>Depressiveness<br>Connectedness<br>Body image<br>Self-efficacy<br>Self-esteem<br>Coping<br>Self-control<br>Sense of coherence |
|  |  | Happiness<br>Life purpose<br>Self-harm<br>Suicidal behaviour<br>Resilience<br>Substance misuse<br>Risky behaviour |
| Social environment | Social environment levels important for adolescents' gender norms | Individual level<br>Interrelational level<br>Community level<br>National level<br>Global level |
|  | Social environment actors that shape the gender norms of adolescents | School environment (teachers, classmates etc.)<br>Media (e.g. television, newspapers, movies)<br>Social media (e.g. Tik-tok, Instagram)<br>Peers/friends<br>Family<br>Sport groups<br>Faith groups<br>Healthcare providers<br>Political parties<br>Law enforcement<br>Civil society<br>Non-Profit-Organisations |
|  | Individual competencies of adolescents to navigate gender norms | Coping skills<br>Agency<br>Navigation<br>Interpersonal relationship skills<br>Critical reflection skills<br>Mental health literacy<br>Respect and empathy for others |

#### Questionnaire

The final questionnaire consists of three main sections and a fourth section about the personal characteristics of the participants. Questionnaire items include content-related closed questions as well as open comment questions and open questions. The sections are briefly presented next. The first section on gender (9 questions) introduces definitions of the key concepts which are sex, gender, and gender norms. The definitions are supposed to ensure that the framework will be based on a shared understanding that challenges a binary understanding of sex/gender. It focuses on assessing which aspects of the gender self-concept (identity) at the individual level are important. Various gender theories or approaches are proposed to be incorporated into the framework and must be rated by the experts by importance. Further, we ask which gender norms are essential that particularly influence adolescents and their mental health.

The section on the mental health of adolescents (4 questions) looks at which mental health outcomes best capture the impact of gender norms on adolescent mental health. We decided not to include mental health disorders as outcomes because this would mean that we would also have to include symptoms as well, which is not the aim of the research. Therefore, several mental health outcomes must be rated by the level of importance, and it must be stated if they can be linked to gender norms and the social environment of adolescents.

In the social environment section (9 questions), we propose five different social environmental levels in an illustration. In addition, we propose actors that may play a role in those social environmental levels for the adolescents and their gendered socialisation process and/or their mental health. These actors should be assigned to the social environment levels in which they play the greatest role. Furthermore, various competencies necessary for adolescents’ navigation with their environment and its social and gender norms are proposed and should be rated by the level of importance.

The last section deals with the experts’ characteristics (10 questions). These encompass the year of birth, the country in which the expert had the most working experience, the country in which the expert currently lives and in which professional environment they are primarily situated (research or policy etc.). Furthermore, we ask experts to rate their topic expertise, meaning how confident they are in the domains of gender or gender norms, the social environment, or the mental health of adolescents to statistically weigh the answers for those with a high competence on the topic. In addition, we assess their sex assigned at birth, the current sex/gender identity, and the sexual orientation of the experts. These characteristics are important to gather to understand their response behaviour in the gender section. Ultimately, we asked the experts to evaluate the questionnaire. An additional document file shows the complete questionnaire in more detail [see Supplementary file 1].

## Discussion

We sought to demonstrate how the Delphi technique can be used in an interdisciplinary public health topic for framework development. We illustrate this with the development of an initial questionnaire on gender norms, the social environment, and the mental health of adolescents for the first Delphi round. The questionnaire development is often seen as the biggest challenge when using the Delphi technique (28).

To ensure that the questionnaire meets methodological standards and adheres to guidelines (18, 29, 17), we developed the questionnaire through a literature review, and we tested for content validity through cognitive interviews, a Delphi expert assessment and group discussions. The validity checks ensure that the questionnaire is more likely to generate more targeted responses to ultimately develop a better conceptual framework. So far, questionnaires for Delphi surveys are typically only tested with a standard pretest like that of the respective online tools (28). Furthermore, we involved international experts from different disciplines in the development process to reflect these characteristics at this early stage.

We encountered several challenges during the development and validation process. They all stem from the characteristics of the resulting framework. The framework must conceptually link gender (norms), the social environment and the mental health of adolescents for quantitative research. For that, it needs appropriate social environment levels that include relevant actors or stakeholders who are carriers of gender norms for or contribute to gender socialisation in adolescents. In addition, it must conceptualise gender and provide a categorisation of gender norms that are relevant to adolescents. Moreover, it needs mental health outcomes that are related to gender and gender norms so that effects and influences can be measured.

The first challenge was the interdisciplinarity of the questionnaire. The goal was to develop a consistent framework which, at the same time, integrates different perspectives and concepts. The experts have different perspectives and priorities and operate with different definitions for terms. Thus, relevant concepts or parts of the concepts and definitions must be adapted and combined so that each participant can follow the logic of the questionnaire items. Moreover, the concepts must be presented in a way that leads to meaningful answer options. The questionnaire items may need to ask for an aspect in a slightly different way or different answer options may need to be provided than in the retrieved frameworks from the literature review. One major decision was that the experts should complete the entire questionnaire and not just the section that relates to their area of expertise. As a result, the experts have to answer questions without expert knowledge in these fields, however, this ensures that the intersections between the topics are covered. The statistical weighing or cross-tabulation of the answers in higher and lower competence is intended to redress this issue. Questions on participants’ self-assessed expertise are also missing in many Delphi surveys (28). Moreover, pop-up information boxes that contain definitions of specific terms are used to inform experts without expertise in that field so that they can answer the question. It is important to emphasize in the questionnaire that the definitions of terms are only examples, so experts with high expertise will not fill in the answer differently simply because of a slightly different understanding of the provided definition. We used cognitive interviews and group discussions to ensure understandability for experts from all included disciplines. Moreover, the introduction of comment boxes as the qualitative part of the questionnaire enables a more concrete understanding of the specific views and ratings of the experts.

The second challenge was the cognitive load of the questionnaire. The objective of the questionnaire involves complex and theoretical questions. We needed to limit the cognitive load to address the generally high drop-out rate in Delphi surveys. To address this challenge, we introduced ranking and rating scales since their benefit lies in a relatively low cognitive and time load (30). This should help the experts to reduce the number of possible answers to the most important and influential aspects. To avoid a “response set” where participants always tick the same answer, we also limited the amount of rating or ranking scales. Besides the introduction of those scales, it was necessary to set some assumptions to reduce the number of questionnaire items. For that, it was particularly useful to predetermine specific aspects, such as the definitions of essential terms, the inclusion of relevant aspects or a limitation to most relevant answer options. Pop-up information boxes also contribute to saving time for the experts.

The third challenge was the quantitative operationalizability of the framework. This requires clear definitions of terms and concepts, clear relationships between them as well as a systematic composition and clear and not too broad questionnaire items. We therefore disentangled the different and often very similar parts of the theoretical concepts and provided answer options (and definitions) that are distinguishable from each other. Another measure was to preselect the most essential answer options and to constantly remind the experts that we were only asking for the most relevant aspects of the topic under investigation. The exclusion of less important aspects can lead to experts mentioning some of the aspects not listed. However, fewer less relevant aspects will likely be mentioned in this way.

### Limitations

An important limitation of this questionnaire development is that we did not statistically test the questionnaire on reliability and validity since the aim was not to develop a scale or a measurement instrument that quantifies well-defined concepts. Another limitation is that we did not execute the literature review in the form of a scoping or systematic review. Furthermore, it remains possible that underlying assumptions about language and interpretations of statement wording are not shared between researchers and respondents. The closed questions may also restrict the depth of participant response and thus the quality of data collected may be diminished or incomplete. However, it was a necessary means to adapt the questionnaire to quantitative research objectives.

## Conclusion

Many Delphi studies do not report on essential elements of the Delphi technique, in particular the specific development steps and the exact questionnaire (5). We have addressed this gap by demonstrating how the Delphi technique can be used to develop a quantitative framework integrating three different public health-relevant topics and by providing the questionnaire of the first Delphi round. Researchers need to preprocess the content and disentangle the theoretical concepts used in their Delphi research to make them accessible to different levels of knowledge, reduce the complexity, provide enough structure and clarity and, if needed, shorten the questionnaire. It should be assessed if researchers from different disciplines should be included in the development process to address challenges at an early stage. Future research should focus on developing methodological guidelines and testing their applicability for different objectives of the Delphi technique (e.g. framework development, interdisciplinary research).

## Data Availability

Data sharing is not applicable to this article as no datasets were generated or analysed during the current study.

## Declarations

### Ethics approval and consent to participate

Ethical approval was obtained from the ethics board at Bielefeld University, Germany, in May 2023 (application number: 2023-111 of 2023/05/22).

### Consent for publication

Not applicable

### Author’s contributions

AA: development of study idea, investigation, execution of cognitive interviews, writing – original draft. EF: participation in group discussion as expert, review and editing. OR: review and editing, CM: supervision of study project, review and editing. All authors read and approved the final manuscript.

## Acknowledgements

We would like to acknowledge and thank Prof. Dr. Michael Häder for contributing his experience with Delphi studies through his evaluation and recommendations for the development of the questionnaire. Special thanks also to Stephanie Batram-Zaantvoort, who participated in the group discussions and contributed her knowledge in the fields of gender, sociology and public health. Further, we would like to acknowledge Diana Podar and Dr. Philipp Jaehn for participating in the cognitive interviews.

## References

1. Mahajan V, Linstone HA, Turoff M. The Delphi Method: Techniques and Applications. Journal of Marketing Research 1976; 13(3):317.

2. Niederberger M, Spranger J. Delphi Technique in Health Sciences: A Map. Frontiers in Public Health 2020; 8:457. Available from: URL: https://www.ncbi.nlm.nih.gov/pmc/articles/PMC7536299/.

3. Nair R, Aggarwal R, Khanna D. Methods of formal consensus in classification/diagnostic criteria and guideline development. Semin Arthritis Rheum 2011; 41(2):95–105.

4. Brady SR. Utilizing and Adapting the Delphi Method for Use in Qualitative Research. International Journal of Qualitative Methods 2015; 14(5):160940691562138.

5. Boulkedid R, Abdoul H, Loustau M, Sibony O, Alberti C. Using and reporting the Delphi method for selecting healthcare quality indicators: a systematic review. PLoS One 2011; 6(6):e20476. Available from: URL: https://journals.plos.org/plosone/article?id=10.1371/journal.pone.0020476.

6. Kågesten A, Gibbs S, Blum RW, Moreau C, Chandra-Mouli V, Herbert A et al. Understanding Factors that Shape Gender Attitudes in Early Adolescence Globally: A Mixed-Methods Systematic Review. PLoS One 2016; 11(6):e0157805.

7. Ryle R. Questioning Gender: A Sociological Exploration. SAGE Publications; 2011. Available from: URL: https://books.google.com.na/books?id=CHHz_p-j9hMC.

8. Martin CL, Ruble DN. Patterns of gender development. Annual review of psychology 2010; 61:353–81. Available from: URL: https://www.ncbi.nlm.nih.gov/pmc/articles/PMC3747736/.

9. Kågesten A, van Reeuwijk M. Healthy sexuality development in adolescence: proposing a competency-based framework to inform programmes and research. Sexual and reproductive health matters 2021; 29(1):1996116. Available from: URL: https://pubmed.ncbi.nlm.nih.gov/34937528/.

10. Johns Hopkins University. Jhpiego: Gender Analysis Toolkit; 2016. Available from: URL: https://gender.jhpiego.org/docs/Jhpiego-Gender-Analysis-Toolkit-for-Health-Systems.pdf.

11. Cislaghi B, Heise L. Using social norms theory for health promotion in low-income countries. Health Promot Int 2019; 34(3):616–23. Available from: URL: https://academic.oup.com/heapro/article/34/3/616/4951539.

12. Bolte G, Jacke K, Groth K, Kraus U, Dandolo L, Fiedel L et al. Integrating Sex/Gender into Environmental Health Research: Development of a Conceptual Framework. International journal of environmental research and public health 2021; 18(22):12118. Available from: URL: https://www.mdpi.com/1660-4601/18/22/12118.

13. Ridgeway CL, Correll SJ. Unpacking the Gender System. Gender & Society 2004; 18(4):510–31.

14. García-Carrión R, Villarejo-Carballido B, Villardón-Gallego L. Children and Adolescents Mental Health: A Systematic Review of Interaction-Based Interventions in Schools and Communities. Front Psychol 2019; 10:918. Available from: URL: https://www.ncbi.nlm.nih.gov/pmc/articles/PMC6491840/.

15. Thomson KC, Romaniuk H, Greenwood CJ, Letcher P, Spry E, Macdonald JA et al. Adolescent antecedents of maternal and paternal perinatal depression: a 36-year prospective cohort. Psychol Med 2021; 51(12):2126–33.

16. Mental health of adolescents; 2021. Available from: URL: https://www.who.int/news-room/fact-sheets/detail/adolescent-mental-health.

17. Moosbrugger H, editor. Testtheorie und Fragebogenkonstruktion: Mit 66 Abbildung und 41 Tabellen. 2., aktual. und überarb. Aufl. Berlin: Springer; 2012. (Springer-Lehrbuch).

18. Tsang S, Royse CF, Terkawi AS. Guidelines for developing, translating, and validating a questionnaire in perioperative and pain medicine. Saudi Journal of Anaesthesia 2017; 11(Suppl 1):S80–S89. Available from: URL: https://www.ncbi.nlm.nih.gov/pmc/articles/PMC5463570/.

19. Svanemyr J, Amin A, Robles OJ, Greene ME. Creating an enabling environment for adolescent sexual and reproductive health: a framework and promising approaches. The Journal of adolescent health : official publication of the Society for Adolescent Medicine 2015; 56(1 Suppl):S7–14.

20. GAGE Conceptual Framework; n.d. Available from: URL: https://www.gage.odi.org/research/conceptual-framework/.

21. Neetu JA, Stoebenau K, Ritter S, Edmeades J, Balvin N. Gender Socialization during Adolescence in Low- and Middle-Income Countries: Conceptualization, influences and outcomes; 2017. Available from: URL: https://www.unicef-irc.org/publications/pdf/IDP_2017_01.pdf.

22. George AS, Amin A, Abreu Lopes CM de, Ravindran TKS. Structural determinants of gender inequality: why they matter for adolescent girls’ sexual and reproductive health. BMJ 2020:19–23. Available from: URL: https://www.bmj.com/content/bmj/368/bmj.l6985.full.pdf.

23. Pulerwitz J, Blum R, Cislaghi B, Costenbader E, Harper C, Heise L et al. Proposing a Conceptual Framework to Address Social Norms That Influence Adolescent Sexual and Reproductive Health. The Journal of adolescent health : official publication of the Society for Adolescent Medicine 2019; 64(4S):S7–S9. Available from: URL: https://www.sciencedirect.com/science/article/pii/S1054139X19300564.

24. Hennegan J, Shannon AK, Rubli J, Schwab KJ, Melendez-Torres GJ. Women’s and girls’ experiences of menstruation in low- and middle-income countries: A systematic review and qualitative metasynthesis. PLoS medicine 2019; 16(5):e1002803. Available from: URL: https://pubmed.ncbi.nlm.nih.gov/31095568/.

25. Kennedy E, Binder G, Humphries-Waa K, Tidhar T, Cini K, Comrie-Thomson L et al. Gender inequalities in health and wellbeing across the first two decades of life: an analysis of 40 low-income and middle-income countries in the Asia-Pacific region. The Lancet Global Health 2020; 8(12):e1473–e1488. Available from: URL: https://pubmed.ncbi.nlm.nih.gov/33091371/.

26. Hammarström A, Johansson K, Annandale E, Ahlgren C, Aléx L, Christianson M et al. Central gender theoretical concepts in health research: the state of the art. Journal of Epidemiology and Community Health 2014; 68(2):185–90. Available from: URL: https://pubmed.ncbi.nlm.nih.gov/24265394/.

27. Hammarström A, Hensing G. How gender theories are used in contemporary public health research. Int J Equity Health 2018; 17(1):34. Available from: URL: https://equityhealthj.biomedcentral.com/articles/10.1186/s12939-017-0712-x.

28. Tietze N, Sterl S, Gerhold L. Delphi-Verfahren in der Praxis: Erfahrungen zum Einsatz von Delphi-Verfahren in den Sozial-und Gesundheitswissenschaften; 2022. Available from: URL: https://refubium.fu-berlin.de/handle/fub188/33999.

29. Tilakasiri KK. Development of New Frameworks, Standards and Principles via Delphi Data Collection Method 2013:1189–94.

30. Niederberger M, Renn O. Das Gruppendelphi-Verfahren: Vom Konzept bis zur Anwendung. Wiesbaden: Springer Fachmedien Wiesbaden; 2018.

