## Supplementary Material File 1 for "A questionnaire for a conceptual framework and interdisciplinary public health research using the Delphi technique – development and validation"

*Supplementary Table 1: Questionnaire for the first Delphi round with section A on gender and gender norms of adolescents, section B on the mental health of adolescents, section C on the social environment and adolescents' competencies and section D on the sociodemographic characteristics of participants*

| **Section A: Gender and gender norms of adolescents** | |
|---|---|
| VV-ID | ID-Variable |
| VV-EW | **[Always display]**<br>Consent to participation / Consent to privacy policy |
| ID | **[Always display]**<br>Pseudonymization variable<br>To identify the participants over the three Delphi rounds, we would like to introduce a 4-digit acronym. Please enter the first two letters of your mother's name plus the two last numbers of your year of birth. |
| A1 | **[Always display]**<br>In addition to gender identity and sex assigned at birth, which of those other aspects of common gender concepts, theories or approaches do you think are most essential for the social environment [The social environment includes the groups to which we belong, the neighbourhoods in which we live, the organization of our workplaces, and the policies we create to order our lives. The physical and social environments do not exist independently of each other; any environment is the result of the continuing interaction between natural and man-made components, social processes, and the relationships between individuals and groups] and the mental health of adolescents? Please put the following aspects in order of importance, putting the aspects that seem most important at the top. If you cannot assign some aspects, leave them unordered.<br>1: Current sex/gender identity [Gender identity is the way you personally identify your gender, such as trans, male/masculine, non-binary, female/feminine, cisgender]<br>2: Sex/gender roles [Gender roles are culturally and socially defined ways that gender is expected to be performed and can often be harmful to people who fall outside of the binary system often upheld through systemic and cultural institutions. Gender roles are often linked to the sex individuals are assigned at birth. Thus, gender roles often reveal the ways that we make culturally contextual assumptions about what different types of bodies should 'do']<br>3: Sex/gender expression [Gender expression is the way a person presents their gender to the world through clothing, dress, physical attributes, mannerisms, behaviours, for instance, masculinized behaviour with feminine expression]<br>4: Sex/gender relations [Gender relations are a specific subset of social relations uniting women and men as social groups in a particular community, including how power – and access to/control over resources – is distributed between the sexes. Gender relations intersect with all other influences on social relations – age, ethnicity, race, religion, etc. – to determine the position and identity of people in a social group]<br>5: Sexuality [Sexuality is a personal identifier that best describes who a person may be attracted to sexually, emotionally, intellectually, or romantically at that current period of time. Some common terms that are associated with sexuality are lesbian, gay, bisexual, pansexual, queer, and many different identities as well] |
| A1-1 | **[Always display]**<br>If you wish, please comment on your rating!<br>1: [free text box]<br>2: No indication |
| A1-2 | **[Always display]**<br>Do you know of any other gender concepts relevant to the social environment and mental health of adolescents?<br>1: No<br>2: Yes [free text box] |
| A2 | **[Always display]**<br>How important do you find these gender approaches to adolescent mental health? Please rate each approach individually.<br>A: Gender continuum: [Considering gender as a continuum between masculinity and femininity]<br>0: Not at all important<br>1: Slightly important<br>2: Important<br>3: Fairly important |

| | |
|---|---|
| | 4: Very important<br>B: Multidimensionality approach: [Gender is multidimensional because it includes characteristics, norms, stereotypes, roles, responsibilities, activities, etc.]<br>0: Not at all important<br>1: Slightly important<br>2: Important<br>3: Fairly important<br>4: Very important<br>Multi-level approach: [Gender is multi-level because it is not only experienced at the individual level but is also imposed by society at different structural levels, e.g. at the institutional or national level that can interact with each other]<br>0: Not at all important<br>1: Slightly important<br>2: Important<br>3: Fairly important<br>4: Very important<br>Intersectionality approach: [Gender intersects with other drivers of inequalities, discrimination, marginalization, and social exclusion, e.g. ethnicity, class, socio-economic status, disability, age, sexual orientation, caste domination etc.]<br>0: Not at all important<br>1: Slightly important<br>2: Important<br>3: Fairly important<br>4: Very important<br>Gender power relations: [Gender entails different power relations, meaning the ways in which gender shapes the distributions of power at all levels of society]<br>0: Not at all important<br>1: Slightly important<br>2: Important<br>3: Fairly important<br>4: Very important<br>Embodiment approach: [Refers to how we, like any living organism, literally incorporate, biologically, the world in which we live, including our societal and ecological circumstances]<br>0: Not at all important<br>1: Slightly important<br>2: Important<br>3: Fairly important<br>4: Very important<br>Decolonial lens: [A decolonial lens is a transformative and liberatory movement that con-siders the effects of imperialism, slavery, racism, and colonialism on directly or indirectly colonised popu-lations, and aims to restructure power imbalances within the fields of global health to establish equitable, mutually beneficial, and reciprocal partnerships be-tween those who continue to profit from the above forces of oppression and those who continue to lose from them]<br>0: Not at all important<br>1: Slightly important<br>2: Important<br>3: Fairly important<br>4: Very important |
| A2-1 | **[Always display]**<br>If you wish, please comment on your rating!<br>1: [free text box]<br>2: No indication |
| A2-2 | **[Always display]**<br>Do you know of any other gender approaches relevant to the social environment and mental health of adolescents?<br>1: No<br>2: Yes [free text box] |

| | |
|---|---|
| | We define gender norms as follows:<br>Gender Norms are produced through social institutions (such as families, schools), social interactions (such as between romantic partners or family members), and wider cultural products (such as books, literature, film and video games). Gender norms refer to social and cultural attitudes and expectations about which behaviours, preferences, products, professions or knowledges are appropriate for female, male and gender-diverse individuals. They draw upon and reinforce gender stereotypes and may be reinforced by unequal distribution of resources and discrimination in the workplace, families and other institutions and. They are constantly in flux as they change by historical era, culture, social contexts or location. |
| A3 | What gender norms can you think of that particularly influence adolescents? Please write your ideas into the text box. E.g. not sharing emotions or weaknesses for adolescent boys<br>1: [free text box] |
| A4 | Which gender norms do you think are of particular importance for the mental health of adolescents? Please write your ideas for gender norms and their possible impact on the mental health of adolescents in the text box. You can also repeat those you mentioned in the previous question. E.g. not sharing emotions can lead to substance misuse.<br>1: [free text box] |
| A5 | Is there something you would like to add that has not been addressed in this part or is there something you would like to comment on?<br>1: No<br>2: Yes [free text box] |
| **Section B: Mental health of adolescents** | |
| B1 | What aspects of adolescent mental health are particularly affected by gender norms? Please rate the importance of the following outcomes for adolescents' mental health according to how much they are affected by gender norms.<br>A: Mental, social and physical well-being<br>0: Not at all important<br>1: Slightly important<br>2: Important<br>3: Fairly important<br>4: Very important<br>B: Depressiveness<br>0: Not at all important<br>1: Slightly important<br>2: Important<br>3: Fairly important<br>4: Very important<br>C: Connectedness<br>0: Not at all important<br>1: Slightly important<br>2: Important<br>3: Fairly important<br>4: Very important<br>D: Body image<br>0: Not at all important<br>1: Slightly important<br>2: Important<br>3: Fairly important<br>4: Very important<br>E: Self-efficacy<br>0: Not at all important<br>1: Slightly important<br>2: Important<br>3: Fairly important<br>4: Very important<br>F: Self-esteem<br>0: Not at all important<br>1: Slightly important |

| | |
|---|---|
| | 2: Important<br>3: Fairly important<br>4: Very important<br>G: Coping<br>0: Not at all important<br>1: Slightly important<br>2: Important<br>3: Fairly important<br>4: Very important<br>H: Self-control<br>0: Not at all important<br>1: Slightly important<br>2: Important<br>3: Fairly important<br>4: Very important<br>I: Sense of coherence<br>0: Not at all important<br>1: Slightly important<br>2: Important<br>3: Fairly important<br>4: Very important<br>J: Happiness<br>0: Not at all important<br>1: Slightly important<br>2: Important<br>3: Fairly important<br>4: Very important<br>K: Life purpose<br>0: Not at all important<br>1: Slightly important<br>2: Important<br>3: Fairly important<br>4: Very important<br>L: Self-harm<br>0: Not at all important<br>1: Slightly important<br>2: Important<br>3: Fairly important<br>4: Very important<br>M: Suicidal behaviour<br>0: Not at all important<br>1: Slightly important<br>2: Important<br>3: Fairly important<br>4: Very important<br>N: Resilience<br>0: Not at all important<br>1: Slightly important<br>2: Important<br>3: Fairly important<br>4: Very important<br>O: Substance misuse<br>0: Not at all important<br>1: Slightly important<br>2: Important<br>3: Fairly important |

| | |
|---|---|
| | 4: Very important<br>P: Risky behaviour<br>0: Not at all important<br>1: Slightly important<br>2: Important<br>3: Fairly important<br>4: Very important |
| B1-1 | **[Always display]**<br>If you wish, please comment on your rating!<br>1: [free text box]<br>2: No indication |
| B1-2 | Do you know of any other mental health outcomes that are linked to experiences of gender for adolescents?<br>1: No<br>2: Yes [free text box] |
| B2 | Is there something you would like to add that has not been addressed in this part or is there something you would like to comment on?<br>1: No<br>2: Yes [free text box] |
| **Section C: Social environment and adolescents' competencies** | |
| C1 | We propose these five social environmental levels (individual, interrelational, community, national, global) as particularly important for the gender norms of adolescents. [This is an adaptation of an ecological model. Research projects dealing with gender and adolescents use social environmental levels because the social environment shapes adolescent health and contributes to their gender socialization]<br>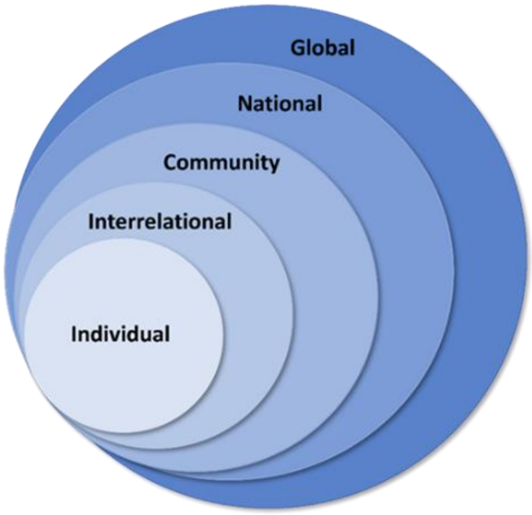<br><br>Do you know of any other social environment levels relevant for experiences of gender for adolescents that are not portrayed in the illustration?<br>1: No<br>2: Yes [free text box] |
| C1-1 | If you wish, please comment on the proposed social environment levels!<br>1: [free text box]<br>2: No indication |
| C2 | In the different proposed social environment levels in the previous question, different actors may play a role in shaping the gender norms of adolescents. Which of the following actors are important to be included in the aforementioned levels? Please choose the level in which they play the greatest role.<br><br>A: School environment [e.g. teachers, classmates etc.]<br>0: Not relevant<br>1: Global level<br>2: National level |

| | |
|---|---|
| | 3: Community level<br>4: Interrelational level<br>5: Individual level<br>B: Media [e.g. television, newspapers, movies]<br>0: Not relevant<br>1: Global level<br>2: National level<br>3: Community level<br>4: Interrelational level<br>5: Individual level<br>C: Social media [e.g. Tik-tok, Instagram]<br>0: Not relevant<br>1: Global level<br>2: National level<br>3: Community level<br>4: Interrelational level<br>5: Individual level<br>D: Friends/Peers<br>0: Not relevant<br>1: Global level<br>2: National level<br>3: Community level<br>4: Interrelational level<br>5: Individual level<br>E: Family<br>0: Not relevant<br>1: Global level<br>2: National level<br>3: Community level<br>4: Interrelational level<br>5: Individual level<br>F: Sport groups<br>0: Not relevant<br>1: Global level<br>2: National level<br>3: Community level<br>4: Interrelational level<br>5: Individual level<br>G: Faith groups<br>0: Not relevant<br>1: Global level<br>2: National level<br>3: Community level<br>4: Interrelational level<br>5: Individual level<br>H: Healthcare providers<br>0: Not relevant<br>1: Global level<br>2: National level<br>3: Community level<br>4: Interrelational level<br>5: Individual level<br>I: Political parties<br>0: Not relevant<br>1: Global level<br>2: National level |

| | |
|---|---|
| | 3: Community level<br>4: Interrelational level<br>5: Individual level<br>J: Law enforcement<br>0: Not relevant<br>1: Global level<br>2: National level<br>3: Community level<br>4: Interrelational level<br>5: Individual level<br>K: Civil society<br>0: Not relevant<br>1: Global level<br>2: National level<br>3: Community level<br>4: Interrelational level<br>5: Individual level<br>L: Non-Profit-Organisations<br>0: Not relevant<br>1: Global level<br>2: National level<br>3: Community level<br>4: Interrelational level<br>5: Individual level |
| C2-1 | If you wish, please comment on the proposed social environment levels!<br>1: [free text box]<br>2: No indication |
| C2-2 | Do you know of any other actors, groups or stakeholders that influence experiences of gender for adolescents?<br>1: No<br>2: Yes |
| C3 | What competencies are necessary for adolescents to navigate gender norms or expectations of their social environment in a way that allows them to develop in a psychologically healthy way?<br><br>A: Coping skills: [Ability to recognise, manage and adapt to different types of internal or external stressors]<br>0: Not at all important<br>1: Slightly important<br>2: Important<br>3: Fairly important<br>4: Very important<br>B: Agency: [e.g. adolescents' ability to realise preferences and choices, express voice and influence and make decisions by drawing on resources at multiple levels]<br>0: Not at all important<br>1: Slightly important<br>2: Important<br>3: Fairly important<br>4: Very important<br>C: Navigation: [How adolescents make decisions where clear-cut right or wrong answers or decisions are not always apparent or available]<br>0: Not at all important<br>1: Slightly important<br>2: Important<br>3: Fairly important<br>4: Very important<br>D: Interpersonal relationship skills: [Ability to communicate and negotiate with others, such as intimate partners, peers, family members and others that influence adolescents' lives]<br>0: Not at all important |

| | |
|---|---|
| | 1: Slightly important<br>2: Important<br>3: Fairly important<br>4: Very important<br>E: Critical reflection skills: [Ability to recognise and understand how social norms and structures shape one's own feelings, behaviours and experiences]<br>0: Not at all important<br>1: Slightly important<br>2: Important<br>3: Fairly important<br>4: Very important<br>F: Mental health literacy: [Ability to gain access to, understand, and use information in ways which promote and maintain good mental health]<br>0: Not at all important<br>1: Slightly important<br>2: Important<br>3: Fairly important<br>4: Very important<br>G: Respect and empathy for others: []<br>0: Not at all important<br>1: Slightly important<br>2: Important<br>3: Fairly important<br>4: Very important |
| C3-1 | If you wish, please comment on the proposed social environment levels!<br>1: [free text box]<br>2: No indication |
| C3-2 | Do you know of any other relevant competencies of adolescents that influence the way they navigate through the expectations of their social environment?<br>1: No<br>2: Yes |
| C4 | Is there something you would like to add that has not been addressed in this part or is there something else you would like to comment on? |
| **Section D: Sociodemographic characteristics of participants** | |
| D1 | In what year were you born?<br>1: [free text box] |
| D2 | In which country did you acquire the most working experience?<br>1: [free text box] |
| D3 | In which country do you live currently? |
| D4 | In which professional environment do you mainly operate?<br>A: Research<br>1: No<br>2: Yes [free text box]<br>B: Development, services and implementation<br>1: No<br>2: Yes [free text box]<br>C: Policy<br>1: No<br>2: Yes [free text box]<br>D: Other<br>1: No<br>2: Yes |
| D4-1 | Please specify in which professional environment you primarily evolve.<br>1: [free text box] |
| D5 | How confident are you in these areas? |

| | |
|---|---|
| | A: Gender<br>0: Does not apply<br>1: Not confident at all<br>2: Slightly confident<br>3: Somewhat confident<br>4: Fairly confident<br>5: Completely confident<br>B: Gender approaches, theory or concepts<br>0: Does not apply<br>1: Not confident at all<br>2: Slightly confident<br>3: Somewhat confident<br>4: Fairly confident<br>5: Completely confident<br>C: Gender (norms) influencing adolescents<br>0: Does not apply<br>1: Not confident at all<br>2: Slightly confident<br>3: Somewhat confident<br>4: Fairly confident<br>5: Completely confident<br>D: Gender socialisation of adolescents<br>0: Does not apply<br>1: Not confident at all<br>2: Slightly confident<br>3: Somewhat confident<br>4: Fairly confident<br>5: Completely confident<br>E: Intersectionality [How gender intersects with other categories and structures]<br>0: Does not apply<br>1: Not confident at all<br>2: Slightly confident<br>3: Somewhat confident<br>4: Fairly confident<br>5: Completely confident<br>F: Mental health<br>0: Does not apply<br>1: Not confident at all<br>2: Slightly confident<br>3: Somewhat confident<br>4: Fairly confident<br>5: Completely confident<br>G: Mental health of adolescents<br>0: Does not apply<br>1: Not confident at all<br>2: Slightly confident<br>3: Somewhat confident<br>4: Fairly confident<br>5: Completely confident<br>H: Other<br>0: Does not apply<br>1: Not confident at all<br>2: Slightly confident<br>3: Somewhat confident<br>4: Fairly confident<br>5: Completely confident |

| | |
|---|---|
| D5-1 | Please specify this other area.<br>1: [free text box] |
| D6 | What sex were you assigned at birth?<br>1: Male<br>2: Female<br>3: Intersex |
| D7 | What is your current sex/gender identity? We ask about gender identity and sexual orientation in this and the following question because it is best practice for surveys and is important for this research topic.<br>1: Female/ woman<br>2: Male/ man<br>3: Trans*/ transman/ transwoman<br>4: Inter*<br>5: Non-binary<br>6: Queer<br>7: An identity not mentioned here<br>8: I do not want to classify as any sex/gender category |
| D8 | What is your sexual orientation?<br>1: Heterosexual<br>2: Lesbian or Gay<br>3: Bisexual<br>4: Or please specify: [free text box]<br>5: Prefer not to say |
| D9 | Is there anything else you would like to share with us?<br>1: [free text box] |
